# The Impact of Late-Career Job Loss and Genotype on Body Mass Index

**DOI:** 10.1101/2020.11.20.20235895

**Authors:** Lauren L. Schmitz, Julia Goodwin, Jiacheng Miao, Qiongshi Lu, Dalton Conley

**Affiliations:** Robert M. La Follette School of Public Affairs, University of Wisconsin-Madison; Department of Sociology, University of Wisconsin-Madison; Department of Biostatistics and Medical Informatics, University of Wisconsin-Madison; Department of Statistics, University of Wisconsin-Madison; Department of Sociology, Princeton University & NBER

## Abstract

Unemployment shocks from the COVID-19 pandemic have reignited concerns over the long-term effects of job loss on population health. Past research has highlighted the corrosive effects of unemployment on health and health behaviors. This study examines whether the effects of job loss on changes in body mass index (BMI) are moderated by genetic predisposition using data from the U.S. Health and Retirement Study (HRS). To improve detection of gene-by-environment (G x E) interplay, we interacted layoffs from business closures—a plausibly exogenous environmental exposure—with whole-genome polygenic scores (PGSs) that capture genetic contributions to both the population mean (mPGS) and variance (vPGS) of BMI. Results show evidence of genetic moderation using a vPGS (as opposed to an mPGS) and indicate genome-wide summary measures of phenotypic plasticity may further our understanding of how environmental stimuli modify the distribution of complex traits in a population.

## Introduction

Recent unemployment shocks from the COVID-19 pandemic have left millions of older workers unemployed. In the U.S. alone, the seasonally adjusted unemployment rate for adults aged 55 and over jumped from 2.6 in February 2020 to as high as 13.6 in April 2020^1^. Recent evidence indicates unemployment rates for workers 55 and older have exceeded those of mid-career workers since the pandemic began—the first time in nearly 50 years that older workers have faced higher unemployment than mid-career workers^2^. For older workers in particular, the scarring effects of unexpected job loss could be severe. Job loss at older ages has been associated with longer periods of unemployment than any other age group^3,4^, higher rates of depression and anxiety^5–7^, and a sharp increase in the need for medical care due to heightened stress levels and gaps in health insurance coverage^8,9^. Further, when reemployed, older workers suffer significant wage penalties and lower levels of employer-offered pension and health insurance^10–12^. All these factors could trigger chronic stress and adverse changes in health and health behaviors^13,14^.

This study expands on past work by examining the degree to which underlying genetic predisposition moderates changes in body mass index (BMI) after a job loss. Genotype-by-environment (G x E) interaction is a fundamental component of population variance for complex traits like BMI, but there has been limited success in identifying G x E effects in human populations due to several challenges, including the complexity of measuring environmental exposures, a need for statistical methods that can screen for genetic effects on phenotypic variability, and inadequate power to detect small G x E effects at loci across the genome^15–17^. To overcome the third challenge of lower power to detect individual effects in the context of multiple hypothesis testing, researchers have used whole-genome polygenic scores (PGSs) constructed from well-powered genome-wide association studies (GWAS) that summarize the genomic contribution to a trait or disease across common variants in the genome^18^. That is, PGSs aggregate thousands of genome-wide genetic influences on a phenotype into a single index using results from GWAS that estimate the association between genetic variants and the conditional mean of a phenotype, which we refer to herein as mGWAS. However, PGSs constructed from mGWAS may not capture the impact of loci that contribute to within-individual variance in an outcome that are more responsive to environmental stimuli (i.e., variance quantitative loci or vQTLs). Since estimating genetic contributions to within-person variability is hindered by a lack of large datasets with genotype data and longitudinal phenotypic data on participants, researchers have developed methods that can detect population-level variance effects that are not driven by mean effects, referred to herein as vGWAS^16,19–23^. In this study, we apply summary statistics from both mGWAS and vGWAS to construct whole genome PGSs for BMI that capture mean effects (mPGS) and variance effects (vPGS)^24^. Evaluating both measures in a G x E framework is necessary because environmental shifts may moderate individuals’ propensity for higher or lower BMI, and/or their propensity towards *changes in BMI* or *BMI plasticity*^21,24^.

Our data come from the U.S. Health and Retirement Study (HRS). The HRS is a nationally representative, longitudinal study with genotype data and over twenty years of sociodemographic data on respondents, including individual-level exposures to involuntary job losses from business closures. We focus specifically on business closures because they are typically the byproduct of external, firm level decisions to restructure or relocate businesses and are therefore considered more exogenous than layoffs or firings, which may be correlated with unobserved health or worker characteristics that could bias G x E estimates ^25–27^. The majority of G x E interaction studies use endogenous measures of the environment that cannot address the non-random distribution of genes across environments. This is important because G x E interactions can, in that case, be proxying a different, unmeasured E that is interacting with G, or G x G interactions (i.e., epistasis) or even E x E (if the measured genes proxy other environments). Specifically, in the case of job loss being endogenous (e.g., for cause), such a measure could be intertwined with a host of unobserved genetic or environmental influences that are associated with health and changes in BMI^28–30^.

To address this, our empirical strategy interacts business closures with, respectively, an mPGS and vPGS in a regression-adjusted semiparametric difference-in-differences (DiD) propensity score matching framework that compares the BMI of those before and after an involuntary job loss with a control group that was not laid off. Combining propensity score matching with DiD estimation makes the model more robust to selection on observables and unobservables with time invariant effects (e.g., ability or worker preferences)^31^. This is necessary because although business closures are plausibly more exogenous than layoffs or firings, it is still possible that workers with unhealthy behaviors or poor health, for example, could select into more vulnerable or volatile industries^32^. To date, we are aware of only one other study that has leveraged a vPGS and a quasi-natural experiment (education reform in the UK) to detect G x E interaction effects on BMI and educational attainment^24^. Results from this study found evidence of mPGS and vPGS interaction effects, indicating that both forms of moderation need to be tested in G x E interaction studies.

In the context of older workers in the U.S., we focus on changes in BMI for two reasons. First, BMI is an inexpensive, non-invasive proxy measure of adiposity that is available for all HRS waves and is predictive of metabolic syndrome and other more difficult to measure anthropomorphic measures like abdominal adiposity that increase risk for cardiovascular disease and type 2 diabetes^33^. In older adults, unintentional weight loss or frailty can also be harmful and indicative of decreased resistance to stressors, resulting in greater vulnerability to disease and disability^34–38^. Past studies have found slight increases in BMI from unemployment^31,39^, particularly in middle-aged workers, but there is no consensus, and some studies that look at the causal impact of job loss on BMI or related health outcomes have been unable to locate an average treatment effect^40–42^. Using the HRS, Salm finds no causal effect of business closures on various measures of physical and mental health, whereas Gallo *et al*. find involuntary job loss is associated with increased depressive symptoms and risk of stroke but not myocardial infarction^6,40,43^. A few studies have explored the possiblity that the health impacts of job loss vary across the distribution of health status^39,42^. For example, using finite mixture models, Deb *et al*. found increases in drinking and BMI were concentrated among workers who were already pursuing unhealthy behaviors pre-job loss, indicating the effects of job loss may be especially problematic for high-risk individuals. However, because genetic data have only recently become available in large population studies, past research was not able to explore the possibility that the impact of job loss on intra-individual fluctuations in BMI may vary across the spectrum of genetic risk.

Second, we focus on changes in BMI because BMI is currently the most well studied phenotypic trait in vGWAS. Previous meta-analyses of mGWAS have identified more than 100 genome-wide significant loci associated with BMI^44–48^. The largest cluster of highly significant genetic variants is located in the *FTO* (fat mass and obesity associated) gene region on chromosome 16. Studies suggest *FTO* polymorphisms increase obesity risk through subtle changes in food intake and preference and affect pathways in the central nervous system that regulate appetite^45,49^. In particular, the SNP rs1421085 underlies the association between the FTO locus and obesity via activation of *IRX3* and *IRX5*, which play a role in the differentiation of adipocyte subtypes^50^. Recent vGWAS have found evidence for loci with variance effects on BMI located in genes responsible for adipocyte differentiation (*PPARG*) and genes implicated in the pathology of obesity, diabetes, atherosclerosis, and cancer (*FTO, PPARG, CCNL1, TCF7L2, ZNF668, GIPR*)^16^. Most BMI-associated loci have their largest impact early in life or during adolescence^51^, although a few loci, which have also been associated with type 2 diabetes or coronary artery disease, exhibit stronger effects in older adults^52^. Past studies have also found genetic moderation of social aspects of the environment that affect BMI, including lifetime socioeconomic status (SES), social norms, birth cohort, and institutional policies^53–55^.

Results indicate that an unexpected job loss did not seem to affect differences in BMI by mPGS; individuals with higher mPGS had higher levels of BMI regardless of whether they were in the treatment or control group. However, we do see suggestive evidence of genetic moderation by vPGS such that individuals in the treatment group varied in their genetic propensity for weight gain or loss, whereas similarly matched individuals in the control group did not. Genetic moderation is particularly pronounced in the lower half of the vPGS distribution; less plastic individuals in the bottom 50% appear to adjust more slowly to environmental changes, resulting in minor weight loss compared to similarly matched individuals in the control group. Results from an event time study analysis show that changes in BMI were detectable up to two years post job loss, but did not persist in the subsequent HRS wave, or up to four years post job loss.

## Results

### Matching quality and summary statistics

**Table 1** displays the means and matching statistics for covariates by treatment group, control group, and matched control group. Detailed variable descriptions can be found in **Supplementary Table 1**, and additional descriptive statistics are reported in **Supplementary Table 2**. After matching, covariates should be balanced with little to no significant differences remaining. We included both the standardized bias and two-sample t-tests for equality of the means to check for significant differences between covariates for both groups (**Methods**)^56^.

**Table 1.** Before-treatment means for treated, control, and matched control groups

|  | Match status | Means |  | %bias | *t*-test |  |
| --- | --- | --- | --- | --- | --- | --- |
|  |  | Treated | Control |  | *t* | *p* > *t* |
| BMI | U | 27.369 | 27.759 | -7.5 | -1.41 | 0.157 |
|  | M |  | 27.575 | -4.0 | -0.55 | 0.582 |
| BMI mPGS | U | 0.030 | -0.001 | 3.0 | 0.59 | 0.557 |
|  | M |  | 0.050 | -2.0 | -0.28 | 0.783 |
| BMI vPGS | U | 0.001 | 0.000 | 0.1 | 0.02 | 0.985 |
|  | M |  | 0.040 | -3.9 | -0.53 | 0.597 |
| Female | U | 0.535 | 0.555 | -4.2 | -0.79 | 0.428 |
|  | M |  | 0.533 | 0.3 | 0.04 | 0.968 |
| Age | U | 57.422 | 57.150 | 6.2 | 1.22 | 0.222 |
|  | M |  | 57.487 | -1.5 | -0.20 | 0.840 |
| Married | U | 0.826 | 0.811 | 4.1 | 0.76 | 0.446 |
|  | M |  | 0.824 | 0.6 | 0.08 | 0.938 |
| No degree | U | 0.107 | 0.059 | 17.5 | 3.84 | <0.0001 |
|  | M |  | 0.095 | 4.3 | 0.53 | 0.595 |
| High school degree | U | 0.666 | 0.517 | 30.5 | 5.66 | <0.0001 |
|  | M |  | 0.618 | 9.8 | 1.36 | 0.173 |
| College degree | U | 0.227 | 0.424 | -42.8 | -7.60 | <0.0001 |
|  | M |  | 0.287 | -13.0 | -1.86 | 0.063 |
| Household income (log) | U | 11.072 | 11.338 | -26.6 | -5.94 | <0.0001 |
|  | M |  | 11.177 | -10.5 | -1.34 | 0.179 |
| Household wealth (\$100k) | U | 3.220 | 3.640 | -4.0 | -0.58 | 0.560 |
|  | M |  | 3.312 | -0.9 | -0.13 | 0.900 |
| Works part time | U | 0.187 | 0.112 | 21.1 | 4.47 | <0.0001 |
|  | M |  | 0.167 | 5.8 | 0.74 | 0.460 |
| Firm size<=4 | U | 0.094 | 0.025 | 29.4 | 8.07 | <0.0001 |
|  | M |  | 0.075 | 8.0 | 0.92 | 0.360 |
| Firm size 5-14 | U | 0.107 | 0.056 | 18.7 | 4.16 | <0.0001 |
|  | M |  | 0.087 | 7.4 | 0.93 | 0.352 |
| Firm size 15-24 | U | 0.056 | 0.026 | 14.9 | 3.46 | <0.0001 |
|  | M |  | 0.044 | 6.2 | 0.77 | 0.439 |
| Firm size 25-99 | U | 0.123 | 0.104 | 5.8 | 1.15 | 0.249 |
|  | M |  | 0.108 | 4.8 | 0.65 | 0.515 |
| Firm size 100-499 | U | 0.126 | 0.189 | -17.5 | -3.10 | 0.002 |
|  | M |  | 0.142 | -4.5 | -0.66 | 0.509 |
| Firm size>=500 | U | 0.382 | 0.584 | -41.1 | -7.77 | <0.0001 |
|  | M |  | 0.425 | -8.8 | -1.20 | 0.230 |
| Agriculture/Fishing/Farming | U | 0.019 | 0.006 | 11.1 | 2.86 | 0.004 |
|  | M |  | 0.009 | 8.7 | 1.14 | 0.255 |
| Construction/Mining | U | 0.075 | 0.032 | 19.3 | 4.61 | <0.0001 |
|  | M |  | 0.065 | 4.3 | 0.51 | 0.610 |
| Manufacturing | U | 0.238 | 0.162 | 19.0 | 3.88 | <0.0001 |
|  | M |  | 0.206 | 8.1 | 1.07 | 0.286 |
| Trade | U | 0.265 | 0.121 | 37.1 | 8.29 | <0.0001 |
|  | M |  | 0.227 | 9.8 | 1.21 | 0.226 |
| Public Services | U | 0.075 | 0.074 | 0.4 | 0.08 | 0.938 |
|  | M |  | 0.066 | 3.3 | 0.47 | 0.639 |
| Finance/Insurance/Real Estate | U | 0.062 | 0.064 | -1.0 | -0.18 | 0.854 |
|  | M |  | 0.063 | -0.5 | -0.06 | 0.950 |
| Public Administration | U | 0.013 | 0.067 | -27.5 | -4.12 | <0.0001 |
|  | M |  | 0.040 | -13.5 | -2.24 | 0.026 |
| Misc. Services | U | 0.251 | 0.470 | -46.6 | -8.35 | <0.0001 |
|  | M |  | 0.321 | -14.8 | -2.10 | 0.036 |
| White Collar | U | 0.642 | 0.713 | -15.3 | -3.00 | 0.003 |
|  | M |  | 0.658 | -3.5 | -0.46 | 0.643 |
| Blue Collar | U | 0.238 | 0.179 | 14.6 | 2.94 | 0.003 |
|  | M |  | 0.218 | 4.8 | 0.64 | 0.526 |
| Service | U | 0.112 | 0.087 | 8.4 | 1.70 | 0.089 |
|  | M |  | 0.112 | 0.0 | 0.00 | 0.996 |
| Job tenure | U | 11.281 | 15.260 | -36.5 | -6.99 | <0.0001 |
|  | M |  | 12.693 | -13.0 | -1.81 | 0.071 |
| Health excellent/very good | U | 0.618 | 0.638 | -4.2 | -0.81 | 0.420 |
|  | M |  | 0.615 | 0.6 | 0.08 | 0.934 |
| Health insurance | U | 0.652 | 0.817 | -37.9 | -8.03 | <0.0001 |
|  | M |  | 0.708 | -12.8 | -1.62 | 0.105 |
| Exercise vigorously 3+ times/week | U | 0.358 | 0.362 | -0.8 | -0.15 | 0.878 |
|  | M |  | 0.360 | -0.3 | -0.04 | 0.967 |
| Ever smoke cigarettes | U | 0.624 | 0.538 | 17.5 | 3.27 | 0.001 |
|  | M |  | 0.576 | 9.7 | 1.33 | 0.184 |
| Cigarettes per day | U | 5.369 | 2.771 | 27.9 | 6.35 | <0.0001 |
|  | M |  | 3.692 | 18.0 | 2.36 | 0.018 |
| Drinks per week | U | 2.305 | 2.353 | -0.9 | -0.16 | 0.871 |
|  | M |  | 2.316 | -0.2 | -0.03 | 0.978 |
| CES-D | U | 0.973 | 0.754 | 14.4 | 2.93 | 0.003 |
|  | M |  | 0.936 | 2.4 | 0.31 | 0.756 |

Abbreviations: U, unmatched; M, matched; CES-D, Center for Epidemiological Studies-Depression 8 item scale. Additional covariates in matching procedure: survey year, regional Census division, additional categories for variables with missing values, and the first 10 principal components of the European ancestry genetic data.

Before matching, individuals affected by a business closure have lower socioeconomic standing, were less likely to have health insurance, and report worse mental health and health behaviors than continuously employed individuals. Labor statistics show they were more likely to work part time, for smaller firms, in the agriculture/fishing/farming, construction/mining, manufacturing, or trade industries, were more likely to be blue collar, and have lower job tenure than workers in the control group. We do not see any significant differences in BMI between treatment and control groups.

After matching, covariates are more balanced overall, and the standardized biases for the majority of variables are at or below 5%, which indicates that mean differences between the treatment and control group are small and the balancing procedure was effective^57^. Notable exceptions include mean differences in education, industry, household income, access to health insurance, and smoking behavior. To minimize any remaining differences between groups, we control for all covariates in our empirical model. Importantly, we do not see any significant difference in the mPGS or vPGS between treatment and control groups before or after matching, indicating the absence of gene-environment correlation (rGE), or evidence of selection into the treatment group by underlying genetic predisposition.

### Construction and predictive performance of Vpgs

Because we are incorporating mPGS and vPGS into our G x E interaction model, it is important to use a vPGS that captures variance effects that are distinct from mean effects^24^. To decorrelate the mean and variance effects, Young *et al*. proposed a dispersion effects test that can identify differences in the variance of the GWAS sample as a whole that are not driven by mean effects at the SNP level^16^. We used dispersion weights from Young *et al*. to construct vPGS in the HRS (**Methods**).

The predictive performance of mPGS for BMI in the HRS and other population-based samples has been well studied ^53–55,58,59^. To evaluate the predictive performance of the vPGS, we fit a Double Generalized Linear Model (DGLM) that allowed us to assess the association between the vPGS and the between-individual variance in BMI in a UK Biobank (UKB) test sample that is independent of our HRS testing sample (**Methods**). **Table 2** reports associations from the DGLM with and without mPGS adjustment (Models 1 and 2, respectively). The dispersion vPGS is significantly associated with the population variance in BMI in the UKB test sample (*p*=1.01E-04), and this association holds after controlling for the mPGS (*p*=1.44E-04).

**Table 2.** Variance polygenic score (vPGS) evaluation in UK Biobank (N=81,375)

|  | **Model 1** |  |  | **Model 2** |  |  |
| --- | --- | --- | --- | --- | --- | --- |
|  | Effect size | SE | *p*-value | Effect size | SE | *p*-value |
| vPGS | 1.93E-02 | 0.005 | 1.01E-04 | 1.89E-02 | 0.005 | 1.44E-04 |

Abbreviations: SE, standard error. Results show the association between the vPGS and the between-individual variance in BMI in a 20% holdout sample of the UK Biobank. Model 2 adjusts for the effect of the mean PGS (mPGS).

### Gene-by-environment (G x E) interaction results

We used a propensity score matched DiD model to evaluate the effect of job loss from a business closure (**Methods**). **Table 3** shows separate propensity score adjusted DiD results from specifications with and without the mPGS and vPGS interactions. Columns 1-2 model changes in BMI without the mPGS and vPGS in the full HRS sample of workers aged 50-70 (1) and in the same-aged European ancestry analytic sample of workers with genotype data (2). Columns 3-4 add the main effect (3) and mPGS interaction effect (4), and Column 5 displays results from the full model with the mPGS and vPGS main effects and interaction effects. All specifications adjust for the conditioning variables used in the propensity score matching that are reported in Table 1. Results without PGSs in Columns 1 and 2 do not condition on genotype in the matching procedure or in the regression analysis. Coefficient *p*-values that pass FDR correction are denoted with an asterisk.

**Table 3.**
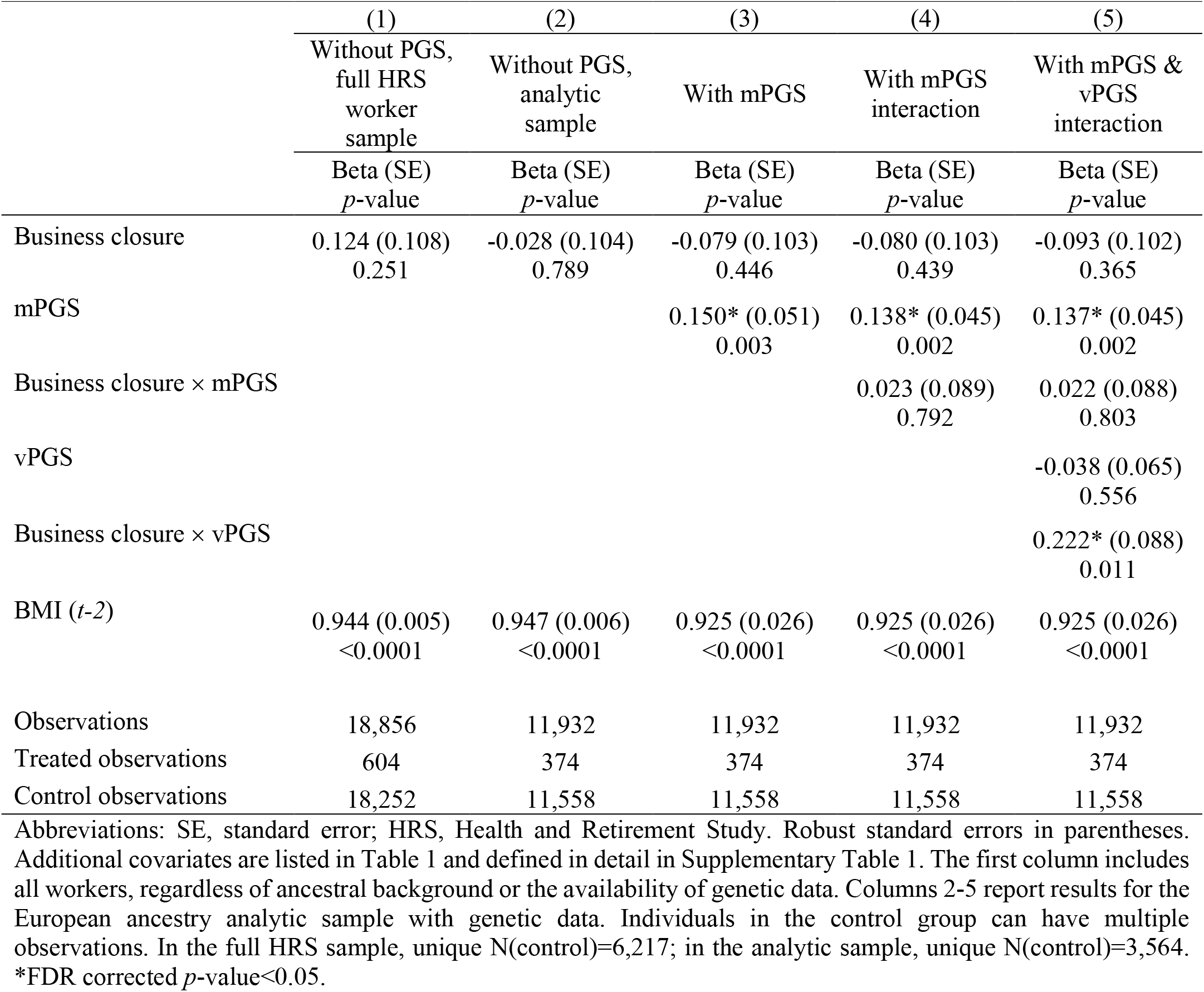
Effect of a job loss from a business closure on BMI, with and without mPGS and vPGS interactions

|  | (1) | (2) | (3) | (4) | (5) |
| --- | --- | --- | --- | --- | --- |
|  | Without PGS,<br>full HRS<br>worker<br>sample | Without PGS,<br>analytic<br>sample | With mPGS | With mPGS<br>interaction | With mPGS &<br>vPGS<br>interaction |
|  | Beta (SE)<br>*p*-value | Beta (SE)<br>*p*-value | Beta (SE)<br>*p*-value | Beta (SE)<br>*p*-value | Beta (SE)<br>*p*-value |
| Business closure | 0.124 (0.108)<br>0.251 | -0.028 (0.104)<br>0.789 | -0.079 (0.103)<br>0.446 | -0.080 (0.103)<br>0.439 | -0.093 (0.102)<br>0.365 |
| mPGS |  |  | 0.150* (0.051)<br>0.003 | 0.138* (0.045)<br>0.002 | 0.137* (0.045)<br>0.002 |
| Business closure × mPGS |  |  |  | 0.023 (0.089)<br>0.792 | 0.022 (0.088)<br>0.803 |
| vPGS |  |  |  |  | -0.038 (0.065)<br>0.556 |
| Business closure × vPGS |  |  |  |  | 0.222* (0.088)<br>0.011 |
| BMI (*t*-2) | 0.944 (0.005)<br><0.0001 | 0.947 (0.006)<br><0.0001 | 0.925 (0.026)<br><0.0001 | 0.925 (0.026)<br><0.0001 | 0.925 (0.026)<br><0.0001 |
| Observations | 18,856 | 11,932 | 11,932 | 11,932 | 11,932 |
| Treated observations | 604 | 374 | 374 | 374 | 374 |
| Control observations | 18,252 | 11,558 | 11,558 | 11,558 | 11,558 |

Abbreviations: SE, standard error; HRS, Health and Retirement Study. Robust standard errors in parentheses. Additional covariates are listed in Table 1 and defined in detail in Supplementary Table 1. The first column includes all workers, regardless of ancestral background or the availability of genetic data. Columns 2-5 report results for the European ancestry analytic sample with genetic data. Individuals in the control group can have multiple observations. In the full HRS sample, unique N(control)=6,217; in the analytic sample, unique N(control)=3,564. \*FDR corrected *p*-value<0.05.

The results in Column 1 are similar in magnitude and direction to the HRS results reported by Deb *et al*, which find a positive, but insignificant main effect from business closures in the full HRS sample on changes in BMI in all ancestry groups^39^. Among genotyped, European ancestry respondents between the ages of 50 and 70 who report being in the labor force, the main effect is still insignificant but negative (Column 2). Columns 3-5 show that the inclusion of both the mPGS and vPGS is necessary to uncover a genetic main effect and an interaction effect: the mPGS captures a significant main effect of genotype on BMI (*p*=0.002), while the vPGS captures a significant G x E effect (*p*=0.011).

Graphically, this can be seen in **Figure 1**, which used estimated parameters from the DiD regression model to predict BMI at different values of the mPGS and vPGS for treated individuals in the wave following a job loss and for corresponding control individuals that were matched to treated individuals in the same HRS wave (**Methods**). In the mPGS figure, which plots predicted BMI based on the mPGS interaction model in Column 3, we see differences in the predicted level of BMI by mPGS, but no significant differences between groups. Conversely, in the vPGS figure, which plots predicted BMI based on the vPGS interaction model in Column 5, there are no differences in the predicted level of BMI by vPGS, but as indicated by the cross-over shape of the interaction, there is suggestive evidence of a G x E interaction, or evidence of environmental moderation by vPGS for the treatment group relative to the control group in the post-treatment wave. Significant differences between treatment and control groups can only be seen in the lower half of the vPGS distribution; individuals below the mean appear more likely to lose weight as a result of a business closure relative to workers with similar vPGS scores. These results seem to indicate that less plastic individuals may adapt more slowly to environmental shifts, at least of the type studied herein.

**Figure 1.**
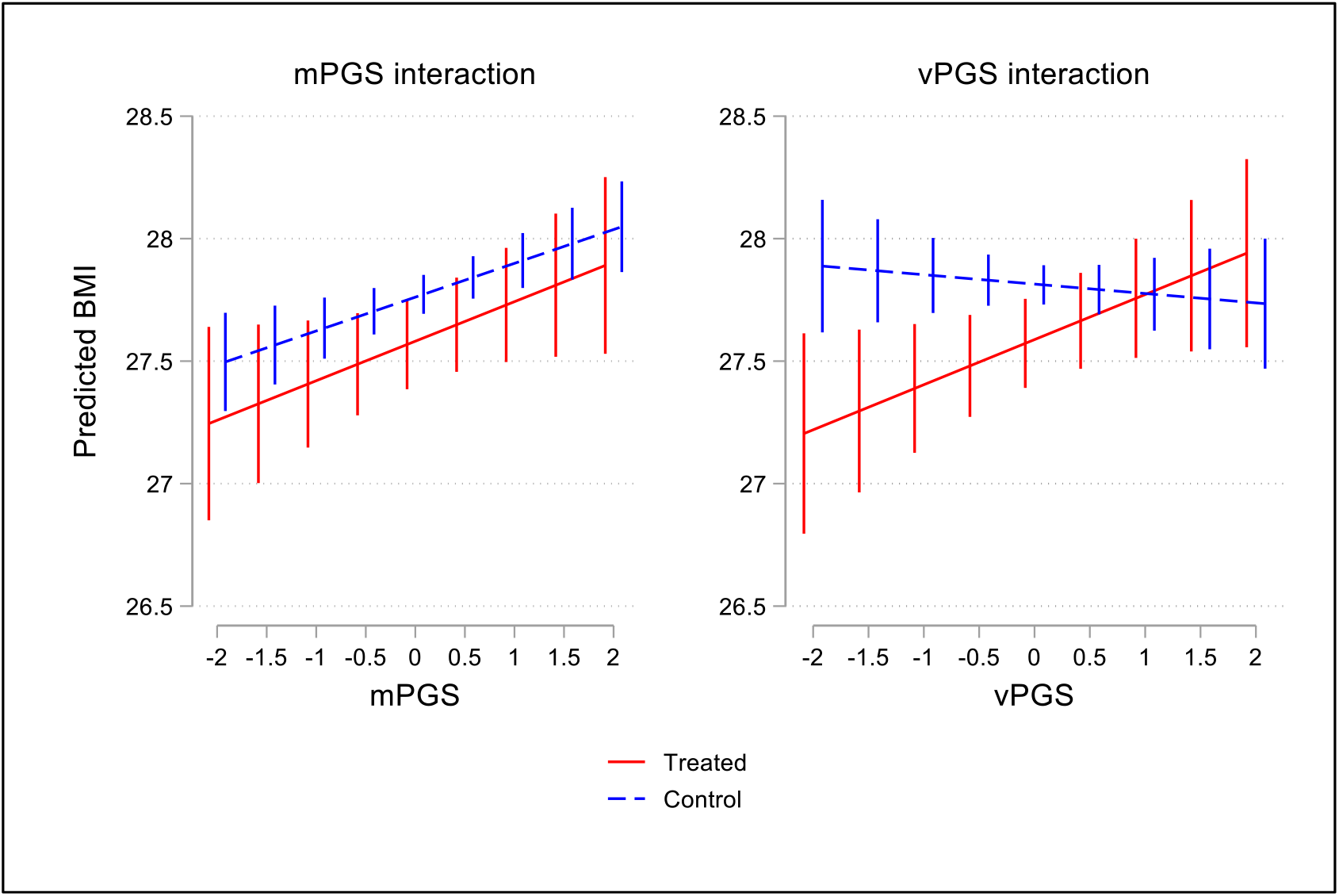
Predicted BMI for treatment and control groups by mPGS and vPGS in the HRS wave following a job loss from a business closure. Figure depicts the predicted BMI for treatment and control groups from the empirical models reported in Table 3 (Column 3) for the mPGS interaction, and Table 3 (Column 5) for the vPGS interaction. Error bars represent 95% confidence intervals.

### Event Time Study

We conducted an event time study (ETS) to assess the validity of our findings and to show the evolution in BMI by vPGS for the treatment and control groups up to four years post job loss (**Methods**). The assumption underlying the DiD research design is that in the absence of an involuntary job loss, BMI would have evolved similarly for the treatment and control groups (i.e., the “parallel trends” assumption). **Figure 2** plots the coefficient estimates from the ETS model, which can be interpreted as the difference in BMI between treatment and control groups (**Supplementary Table 3**). The first panel of Figure 2 indicates the presence of parallel trends in BMI prior to a business closure for the full sample—i.e., the difference between treatment and control groups is close to zero and not statistically significant. We then estimated separate event time study regressions for respondents in the top and bottom 50% of the vPGS distribution to compare trajectories in BMI for treatment and control groups by vPGS. Similar to the results in Figure 1, we found suggestive evidence that individuals in the bottom 50% of the vPGS distribution have a lower BMI on average compared to the control group (*p*=0.035). Effects did not appear to persist in the next HRS interview wave at *t* + 2, or up to four years post job loss.

**Figure 2.**
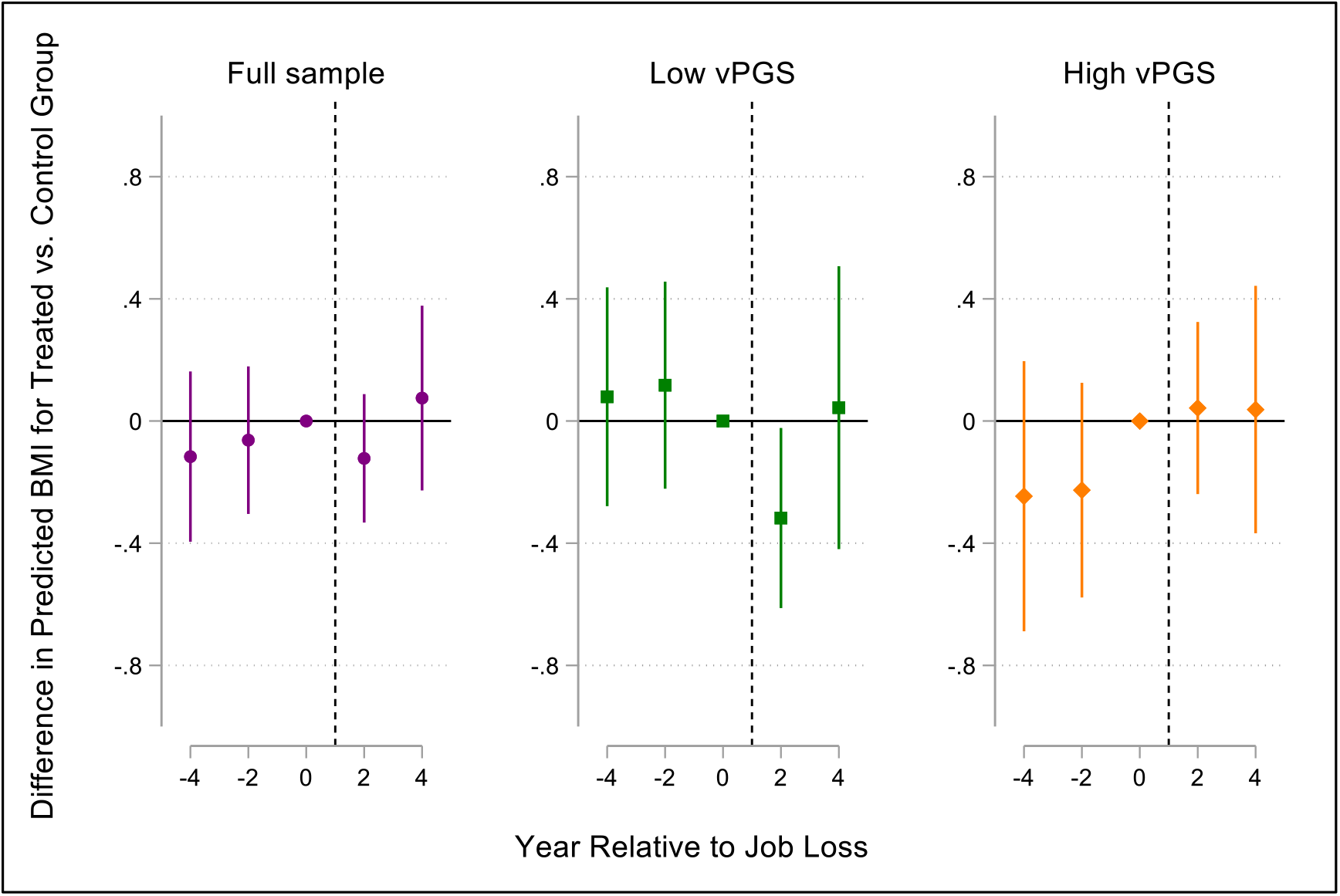
Difference in predicted BMI for treated versus control groups by the year relative to job loss for the full sample and by samples stratified at the vPGS median. The figure plots the coefficients from an event time study model for the full sample and by samples stratified at the vPGS median (**Methods**). Error bars represent 95% confidence intervals. The reference category is BMI in *t*-2, or BMI at baseline before the job loss occurred for treated individuals (represented here as year 0). The dotted line depicts the approximate time point that the job loss occurred—i.e., some point between the baseline year and the current wave. *Date*: This manuscript was compiled on November 20, 2020. *Date*: This manuscript was compiled on November 20, 2020.

## Discussion

Gene-environment interplay is a fundamental biological process that influences the diversity of outcomes we observe in human populations^60^. However, because genetic differences are tightly interwoven with environmental differences, it is challenging to identify genomic and environmental factors underlying phenotypic plasticity. The search for interaction effects is further complicated by the fact that the majority of GWAS methods are unable to separate genetic effects on phenotypic variability from effects on the mean or level of trait values^16^. As a result, most PGS x E interaction studies cannot detect interaction effects that are driven by loci that affect plasticity^24^. Research suggests that SNPs associated with the variance of BMI (vQTLs) are highly enriched for G x E interactions, and that these vQTLs play in an important role in cellular response to the external environment^16,22^.

In this study, we used a natural experiment to investigate whether the effect of job loss on BMI—a stressful and often debilitating lifetime event—is moderated by genetic predisposition. To incorporate genetic effects associated with the population variance in BMI, we followed Johnson *et al*. and used summary statistics from recently developed GWAS methods that can separate SNP mean and variance effects^16^ to construct vPGS for BMI^24^. Using both an mPGS and a vPGS, we find stronger evidence for a vPGS x E interaction. A shift in the environment did not seem to affect differences in BMI by mPGS; individuals with higher mPGSs had higher levels of BMI regardless of whether they were in the treatment or control group. However, we do see differences between the treatment and control groups by vPGS such that less plastic or lower vPGS treated individuals showed signs of weight loss compared to similarly matched control individuals, perhaps because they adjust more slowly to an environmental shock compared to high vPGS individuals. ETS analysis suggests that treatment and control groups evolved similarly prior to treatment, and that findings are detectable up to two years post job loss.

It is important to note that we deployed, by necessity, a vPGS that was constructed from weights that were trained in a discovery sample to predict variation *between* individuals net of mean effects. However, we are using this measure in analysis that examines *within*-subject variation. This is an important distinction that may inform the interpretation of our results. Namely, our theory is that highly plastic individuals are better able to adapt to changing environmental contexts and thus maintain a more stable weight in the face of job loss. We classify individuals as plastic or non-plastic based on their score from a genetic model that predicts whether an individual scores higher on a cross-individual model of dispersion (independent of mean effects). In a sense, this means that someone with a higher vPGS has more “noise” in their prediction than another individual—that is, s/he is less well-predicted from a levels (mean values) regression than someone with a lower score. Someone who is low on the plasticity score has a BMI that is predicted better by their levels effects than someone who is high on the score. This, in turn, we think is indicative of someone whose phenotype is more affected by non-additive genetic effects (i.e., epistasis) as well as by environmental effects. That is, imagine two groups: One with a low vPGS and one with a high vPGS. It is the lower-vPGS group that may have a narrow range around a population mean BMI of 25 (say, SD = 2 units), while those with a high vPGS may display the same mean BMI in their group (25) but have a wider dispersion (SD = 4 units). Thus, high vPGS individuals do a better job of buffering differences in environments they encounter and, as a result, their phenotypes vary more widely. In one sense of “plasticity” they are higher, as their phenotype varies more.

Turning to our within-person analysis in the HRS, low vPGS-scoring individuals may be more “stable” in their weight from year to year, given the smaller “error” term from the levels regression. Indeed, when we simply compare the standard deviation of BMI within individuals in our HRS sample across waves by a quartile split in the vPGS, irrespective of treatment status, we find that individuals in the lowest quartile of the vPGS score range display a (non-significantly) lower within-person standard deviation in BMI than individuals in the highest quartile (**Supplementary Table 4**). Thus, in the absence of a specific, measured environmental shock, an individual with a low vPGS score derived from the cross-person discovery analysis has more phenotypic *stability* in the within-person analysis (i.e., less plasticity). This may be one reason why we are able to detect significant differences between treated and control individuals up to two years post job loss in the lower half of the vPGS distribution—i.e., in the presence of an unexpected job loss, it takes less plastic individuals longer to recover to their pre-job loss weight. On the other hand, it could be that less plastic individuals display a more stable weight trajectory overall— regardless of environmental forces, rendering our ability to see a “specific” environmental effect more clearly. Although our results point to the former explanation, we acknowledge that we cannot draw any definitive conclusions from this study and more analysis is needed to determine the extent to which a vPGS constructed from a cross-person GWAS captures within-person vQTL effects.

Overall, with only 374 individuals in our treatment group, we acknowledge that we are likely underpowered to detect precise G x E effects. This could in part explain why we are unable to detect effects between treatment and control groups in the upper part of the vPGS distribution. Specifically, because vPGS individuals display a higher within-person standard deviation in BMI, it may be harder to detect differences between treatment and control groups in smaller samples because their weight oscillates more between waves, independent of any particular treatment effect, as mentioned above. Conversely, it’s possible that the true shape of the interaction does not display a crossover effect at higher levels of the vPGS—perhaps because of the larger within- or between-person standard deviation in BMI. Either way, due to a lack of detailed job loss data in other population studies that also collect genetic data on participants, we were unable to pursue replication of our quasi-experimental approach in other samples. Thus, we caution that our results are suggestive, and further analysis that can leverage quasi-experimental variation is needed to assess the validity of our approach.

### Strengths and Limitations

There are several limitations of the HRS data, all of which may bias our estimates downwards or reduce the precision of our estimates. First, we only observe BMI in the HRS every two years, which makes it difficult to assess stress-related changes in BMI that are more proximal to the timing of the event, or in the months immediately pre- and post-job loss. It is entirely possible that high vPGS individuals gained or lost more weight than low vPGS individuals in the months following a job loss but they bounced back quicker to their pre-job loss weight than low vPGS individuals, which would make it more difficult to detect differences between high vPGS treatment and control individuals in the subsequent HRS wave. Second, to obtain the largest sample of treated individuals, we were limited to using self-reports of BMI, which may induce measurement error in our estimates. In 2006, the HRS did start collecting in person, objective measures of BMI; however, these measures are only collected at every other wave, or every four years, and are not available for all treated individuals. Third, because the HRS is a sample of older individuals who were genotyped in 2006, 2008, or 2010, our results may be subject to mortality selection. To reduce the potential of mortality selection, we limited our analyses to individuals born after 1930^61^.

In addition, there is significant complexity surrounding obesity and aging such that differences in BMI may not indicate an actual change in body fat. Higher BMI at midlife is a risk factor for age-related disease and early mortality, however at older ages it might be somewhat protective of mortality because age-related diseases and aging itself are wasting conditions that stimulate significant weight loss. Therefore, while incrementally higher BMI in midlife is more likely a measure of risk for disease, later in life it may actually signal the absence of disease. In addition, individuals generally lose muscle mass with increasing chronological age, meaning older individuals could maintain a constant BMI while simultaneously losing lean body mass and gaining a greater portion of adiposity^62^. Thus, any increases in BMI from a job loss may be offset by these other countervailing trends that are inherent to the aging process, which may in part be another explanation the null findings we report for more plastic individuals in the top half of the vPGS distribution. Furthermore, the relatively nominal findings we report may reflect a greater culmination of environmental and lifestyle factors on adiposity in older adults that overwhelm any genetic effects. The genomic influence on BMI has been shown to both weaken over the life course and increase in magnitude since the current obesity epidemic began in the mid-1980s^55,58,59,63^. While performing exact matching on survey year may in part account for differences in the obesogenic environment across cohorts, our sample size of treated individuals is too small to explore more detailed cohort analysis.

Finally, a significant limitation of this study is that were limited to conducting analyses in individuals of European decent. We focus on individuals of European decent because comparable GWAS in other ancestral populations are currently unavailable. Estimates from a European ancestry GWAS are not necessarily accurate or valid in other ancestral populations, and PGSs constructed from European ancestry GWAS summary statistics will not have the same predictive power for individuals from other ancestral backgrounds^64,65^. Restricting our analysis to one ancestral group is also important in that SNPs within regions of interest may tag different causal variants if the underlying linkage disequilibrium (LD) structure varies across ancestral groups^65,66^. Thus, we caution that PGSs constructed from European ancestry GWAS cannot be generalized to other ancestral populations. On the environmental side, limiting our analysis to white HRS respondents restricts the scope of potential job loss effects that we can observe. Race powerfully shapes structural- and institutionally-derived differences in occupational sorting and occupational opportunities across the life course^67,68^. For example, white HRS respondents were more likely to work in higher status jobs with better working conditions than their Black counterparts^69^, and following a job loss, they were more likely to be reemployed or have additional economic resources to buffer stressful declines in income^70–72^, all of which may further bias effects from this study downwards.

These limitations are counterbalanced by several strengths of our study. The use of a large, nationally representative cohort of individuals from the same ancestry group is an advantage in that it both increases our power to detect effects while also minimizing the presence of ascertainment bias and other selection issues. Having access to detailed, longitudinal job loss data in the HRS also allowed us to exploit a quasi-experimental research design that limited the treatment group to individuals who lost their job due to a business closure while also creating a control group that is matched on a rich suite of characteristics pre-job loss. Current G x E interaction studies that utilize population data are often unable to separate gene-environment correlation (rGE) from G x E effects, which limits our understanding of social-environmental effects on health^27^. Finally, to our knowledge, this is one of the first studies to integrate genetic measures that can separately capture phenotypic mean and variance effects into PGS x E interaction analysis.

## Conclusion

Control of phenotypic variability, both within and between individuals, is a fundamental property of biological systems that impacts how species adapt to environmental changes^73–75^. Incorporating vPGS measures into G x E interaction research may further our understanding of how and to what extent environmental stimuli modify the distribution of anthropomorphic traits in a population. In particular, sizable unemployment shocks from the Great Recession and the COVID-19 pandemic have highlighted the importance of understanding the short- and long-run health consequences of business cycles. Future studies that are able to observe the biology underlying these types of large, social-environmental effects on physiological changes that precede disease promises to inform new opportunities for effective intervention^76^.

## Methods

### Standardized bias estimates

The standardized bias compares the distance between the marginal distributions, or the difference in sample means between the treated 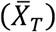 and matched control 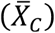 subsamples as a percentage of the square root of the average of the sample variances in both groups for a covariate *X* (Rosenbaum and Rubin, 1985):

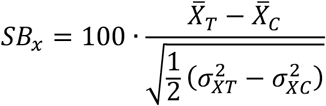

### Health and Retirement Study (HRS) data

The HRS is a nationally representative, longitudinal panel study of individuals over the age of 50 and their spouses that is sponsored by the National Institute on Aging (NIA U01AG009740) and conducted by the University of Michigan^77,78^. Launched in 1992, the HRS introduces a new cohort of participants every six years and interviews around 20,000 participants every two years. To maximize sample size, we compiled data from 13 waves (1992-2016). Information on job loss and smoking behavior was obtained from the 1992-2016 Public Use Core Files; demographic and socioeconomic data came from the RAND Data File (version P).

Genotype data on ∼15,000 HRS participants was collected from a random subset of the ∼26,000 total participants that were selected to participate in enhanced face-to-face interviews and saliva specimen collection for DNA in 2006, 2008, and 2010. We restricted our sample to individuals of European ancestry who were between the ages of 50-70 who reported working part-time or full-time in the previous wave and who were not self-employed. The final sample consists of 3,938 workers with 11,932 observations.

### HRS genotyping and quality control

Genotyping was conducted by the Center for Inherited Disease Research (CIDR) in 2011, 2012, and 2015 (RC2 AG0336495, RC4 AG039029). Full quality control details can be found in the Quality Control Report^79^. Genotype data on over 15,000 HRS participants was obtained using the llumina HumanOmni2.5 BeadChips (HumanOmni2.5-4v1, HumanOmni2.5-8v1), which measures ∼2.4 million SNPs. Genotyping quality control was performed by the Genetics Coordinating Center at the University of Washington, Seattle, WA. Individuals with missing call rates >2%, SNPs with call rates <98%, HWE p-value < 0.0001, chromosomal anomalies, and first-degree relatives in the HRS were removed. Imputation to 1000G Phase I v3 (released March 2012) was performed using SHAPEIT2 followed by IMPUTE2. The worldwide reference panel of all 1,092 samples from the Phase I integrated variant set was used. These imputation analyses were performed and documented by the Genetics Coordinating Center at the University of Washington, Seattle, WA. All positions and names are aligned to build GRCh37/hg19.

Principal component (PC) analysis was performed on a selected set of independent SNPs to identify population group outliers and to provide sample eigenvectors as covariates in the statistical model to adjust for possible population stratification and were provided by the HRS. The European ancestry sample included all respondents that had PC loadings within ± one standard deviations for eigenvectors one and two in the PC analysis of all unrelated study subjects and who self-identified as White on survey data. A second set of principal components was then calculated within the European ancestry sample to further account for any population stratification within the sample. The genotype sample has been defined by the HRS and is available on dbGaP^80^.

### Mean polygenic score (mPGS) construction

We calculated a linear mPGS for the HRS sample based on a GWAS of 457,824 European ancestry individuals in the UK Biobank^48^. Imputed HRS genotype data were accessed through dbGap (phs000428). The mPGS BMI score was constructed in PRSice^81^ by taking a weighted sum across the number of SNPs (*n*) of the number of reference alleles *x* (zero, one, or two) at that SNP multiplied by the effect size for that SNP (β):

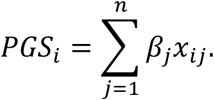

GWAS summary statistics were pruned for linkage disequilibrium (LD) using the clumping procedure in PLINK (R^2^=0.1, range=1000kb)^82,83^. Since these GWAS summary statistics were pre-clumped, no LD-clumping or p-value threshold was implemented in PRSice. After LD clumping was applied, 90,326 SNPs were used to construct the BMI mPGS. The mPGS was standardized to have a mean of zero and a standard deviation of one for all analyses.

### Construction of the variance polygenic score (vPGS)

We calculated BMI vPGS for HRS participants of European ancestry. SNP weights in the vPGS were based on dispersion effects estimated in the UKB using the heteroskedastic linear mixed model (HLMM) approach^16^. Pre-pruned HLMM summary statistics were obtained from Young *et al*.^16^. We did not perform additional LD-clumping or p-value thresholding to filter variants. A total of 242,870 SNPs remained in the vPGS model after overlapping the HLMM summary statistics and HRS genotype data. The vPGS was constructed in PRSice^81^ and standardized to have a mean of zero and a standard deviation of one for all analyses.

### Validation of the variance polygenic score (vPGS)

Performance of the vPGS was assessed using European ancestry UKB samples identified from genetic PCs (data field 22006). To avoid overfitting, HRS samples were not used for model validation. Quality control procedures for the UKB genetic data have been described elsewhere^84^. We excluded participants recommended by UKB (data field 22010), those with conflicting genetically-inferred (data field 22001) and self-reported sex (data field 31), and those who withdrew from the study. We randomly apportioned UKB participants (N=406,873) into training (N=325,498) and testing sets (N=81,375), with an 80-20 split. We applied the HLMM approach to estimate the dispersion effect of each SNP on BMI using samples in the training set, controlling for sex, age, age^2^, age^3^, age × sex, age^2^ × sex, age^3^ × sex, genotyping array, and the first 40 genetic PCs. Following Young *et al*., we analyzed related and unrelated samples in the training set separately and performed fixed-effect meta-analysis to combine the results^16^. Related samples were inferred from genetic kinship (third-degree relatives or higher; data field 22021). Random effects were included to account for genetic relatedness in the analysis of related samples. We then pruned SNPs following Young *et al*. and used dispersion effect estimates to generate vPGS for samples in the testing set.

We then fitted a Double Generalized Linear Model (DGLM) to associate the vPGS with the between-individual BMI variance in testing samples^85^. The DGLM takes the form of

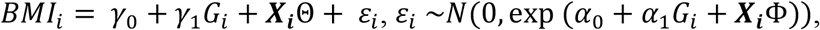

where *BMI*_*i*_ denotes the inverse normal-transformed BMI of individual *i, G*_*i*_ is the vPGS of individual *i*, ***X***_*i*_ is the vector of covariates including sex, age, age^2^, age^3^, age × sex, age^2^ × sex, age^3^ × sex, genotyping array, and the first 40 genetic principal components. Here, α_1_ quantifies the effect of vPGS on the variability of BMI and is the parameter of interest in this analysis. The vPGS was standardized with a mean of 0 and a variance of 1. We fitted DGLM using the dglm package^86^ in R.

To assess the performance of vPGS after adjusting for the effect of mPGS, we performed a standard, mean-effect GWAS of BMI on the training set and used the effect estimates to generate mPGS for the testing samples. GWAS summary statistics were pruned for LD using the clumping procedure in PRSice (R2=0.1, range=250kb) when calculating the mPGS^81^. We then fitted the same DGLM model as above with the mPGS added to the vector of covariates. The mPGS was standardized with a mean of 0 and a variance of 1.

### Treatment and control groups

For each observation, we used information from two waves— before and after treatment. Before treatment (*t*-2), all respondents were working for pay either full- or part-time. At the following HRS interview two years later (*t*), respondents in the treatment group report they were no longer working for their previous-wave employer. These respondents were asked why they left their employer. Possible answers included ‘business closed’, ‘laid off/let go’, ‘poor health/disabled’, ‘quit’ ‘family care’, ‘better job’, ‘retired’, ‘family moved’, ‘strike’, ‘divorce/separation’, ‘transportation/distance to work’, and ‘early retirement incentive/offer’. Respondents could report up to three reasons. Our definition of exogenous job loss includes observations that reported being laid off due to a business closure. We excluded workers who also stated that they quit or left for health reasons but included workers who stated they were also laid off or let go^40^. For the control group, we used individuals who reported working for the same employer the entire time they were in the sample—i.e., we did not include individuals in the control group if they ever quit their job or were laid off for any reason. Treated individuals are only in the analytic sample for two waves, or pre- and post-job loss. Control individuals can be in the analytic sample for multiple HRS waves. In the whole observation period, there were 374 instances of involuntary job loss—i.e., our treatment group consisted of 374 workers who reported being laid off due to a business closure. The control group consisted of 11,558 observations on 3,564 workers.

### Difference-in-differences (DiD) approach

We used DiD estimation combined with nonparametric kernel matching to estimate the average treatment effect on the treated (ATT) by genotype, or the change in BMI by genotype brought about by the job loss of those who actually lost their job^31,87^. This approach compares individuals who have been laid off due to a business closure with a group of similar individuals who are still working for their same employer. To construct a control group with a similar distribution of covariates as the treatment group, the kernel-based matching estimator uses a distance-weighted average of all propensity scores in the control group to construct a counterfactual outcome for each individual in the treatment group. These weights were applied to the DiD regression model to obtain a balanced sample of treated and untreated individuals. The coefficients from the DiD regression were then used to estimate the ATT by mPGS and vPGS.

A traditional DiD setting assumes that after conditioning on a vector of observables *X*, the BMI of individuals in the treatment group would have evolved similarly over time to the BMI of individuals in the control group if they had never been laid off:

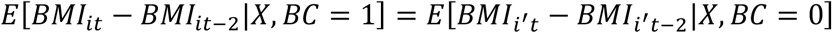

Where *BMI*_*i*t_ − *BMI*_*i*t*−*2,_ refers to the change in BMI before and after the treatment, *BC* denotes the treatment group indicator (i.e., whether an individual lost their job due to a business closure), and *i* ′ denotes an individual in the control group with the same characteristics as individual *i* in the treatment group. While conditioning on genotype and a rich set of covariates minimizes the possibility of violating this assumption, other systematic differences between the treated and control groups may remain even after conditioning on observables.

To minimize potential confounding from unobservable characteristics, we used the weights from propensity score matching (*W*) to reduce unmeasured differences between the treatment and control groups that could bias estimates:

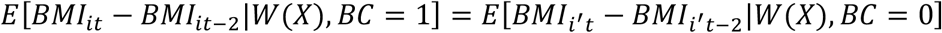

Covariates used to estimate the propensity score, or the probability of treatment, were also included in the DiD regression model. Thus, coefficients from the regression-adjusted semiparametric DiD matching estimator are considered “doubly robust” because the estimator is consistent if the regression model or the propensity score model is correctly specified^88,89^. As a result, the DiD matching estimator accounts for selection on observable and unobservable variables with time invariant effects, or the model allows for systematic differences between treatment and control groups even after conditioning on observables^90^.

### Difference-in-differences (DiD) empirical strategy

Our empirical strategy can be broken down into three parts. First, we estimated propensity scores using a probit regression that regresses business closures on the mPGS and vPGS, as well as a rich set of covariates that are both standard in the job loss literature and satisfy the conditional independence assumption—i.e. they influence job loss and/or changes in BMI^87,91^. In addition, we only conditioned on observables that were unaffected by job loss (or the anticipation of it), or variables that were either fixed over time or measured in *t*-2 ^57^. A complete list of covariates can be found in **Supplementary Table 1**. To avoid losing observations with missing information on a covariate, we set missing values equal to zero and included an additional dichotomous variable that is equal to one if the observation is missing. As a result, matching is not only on observed values but also on the missing data pattern^31,92^. Throughout, we restricted our analysis to the region of common support, or the subset of individuals in the control group that were comparable to individuals in the treatment group^91^. Specifically, we dropped treatment observations whose propensity score was greater than the maximum or less than the minimum propensity score of the controls.

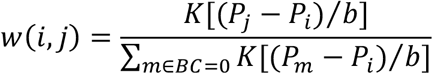

We used the estimates from the probit regression to compute the weights for the control group with kernel matching, a nonparametric matching estimator that uses the weighted averages of all observations on common support to construct the counterfactual outcome^87,90^. Specifically, the weight given to a non-treated individual *j* was in proportion to the closeness of their observables to treated individual *i*:

Where *P* is the propensity score for individual *i* or *j* in the treated or control group, respectively, *K*[⋅] is the kernel function, and *b* is the bandwidth parameter. We used the program psmatch2^93^ in Stata 14 to compute *w*(*i, j*) with the Epanechnikov kernel function and a bandwidth of 0.06^87^. In addition, when computing the weights, we performed exact matching on survey year and sex in *t*-2. This ensured 1) individuals who were laid off were matched with controls from the same time period, and 2) treated individuals were grouped with same-sex non-treated individuals.

In the final step, we incorporated the weights from propensity score matching into the DiD regression model:

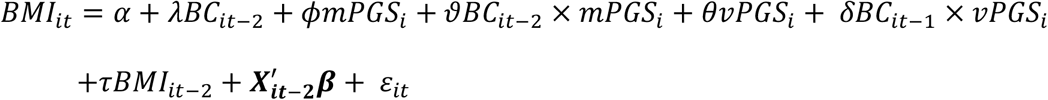

Where *BC* is an indicator for job loss due to a business closing in the years between HRS survey waves, or between *t*-2 and *t* for individual *i*, ***X*** is a vector of observable time invariant and variant covariates measured at *t*-2, including the first 10 principal components of the genetic data. We also include *BMI*_t*−*2,_ to control for baseline BMI, or to estimate deviations in BMI between *t*-2 and *t*. All regressions were estimated with robust standard errors.

### Estimating average treatment effects by genotype

Estimated parameters from the DiD regression model were used to estimate the conditional mean or predicted BMI for treated and untreated individuals at various values of the mPGS and vPGS (**Figure 1**). For example, the BMI for treated (*BC*=1) and untreated (*BC*=0) individuals with hypothetical mPGS and vPGS values at 0 and 1, respectively, would be estimated as follows:

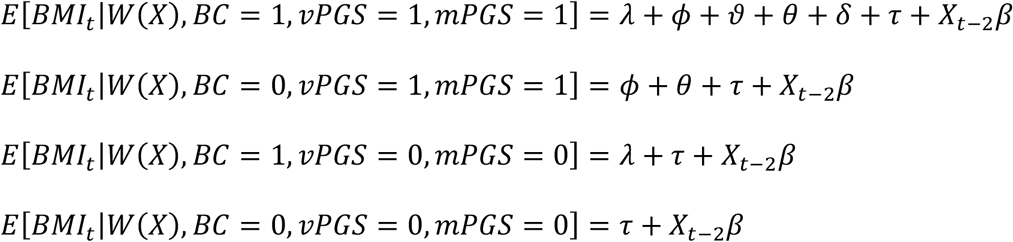

From here, the ATT can be estimated by taking the difference in *E*[*BMI*_t_ |*W*(*X*)] between treated and non-treated individuals with the same mPGS and vPGS values:

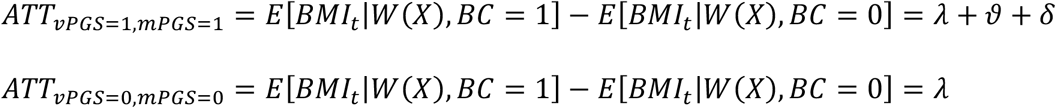

### Event time study analysis

We estimated an event time study (ETS) model for individuals in the top and bottom 50% of the vPGS distribution using the following specification:

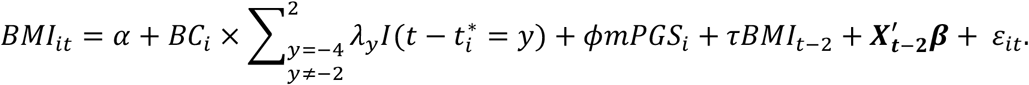

This model is similar to the DiD model outlined above except the business closure term is replaced by a series of event terms that are the product of indicators for each HRS survey year (*y*) relative to the survey year the respondent reported a job loss 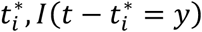, and their treatment status (*BC*_*i*_). The omitted category is the survey year prior to treatment (*y* ≠ −2). We also present ETS results for the full sample that includes controls for the vPGS. Each estimate of λ_*y*_ gives the difference in BMI for treated individuals compared to non-treated individuals relative to this excluded year. If outcomes were evolving similarly for treated and untreated individuals prior to a business closure, the coefficient estimates for *y* < 0 should be close to zero and not statistically significant.

## Data Availability

All phenotype and socioeconomic data are publicly available on the Health and Retirement Study (HRS) website. Genetic data on HRS participants are distributed through the NCBI Database of Genotypes and Phenotypes (dbGaP).

## Acknowledgements

This research was funded by the following awards from the National Institute on Aging (NIA): K99 AG056599, R00 AG056599, P30 AG012846, T32 AG000221 (Schmitz), and P30 AG17266 (Lu). The content is solely the responsibility of the authors and does not necessarily represent the official views of the National Institutes of Health. The authors would like to thank Jason Fletcher, Rebecca Johnson, and Ramina Sotoudeh for helpful comments and feedback on earlier versions of this draft.

## Supplementary Information

**Supplementary Table 1.** Variable definitions

|  |  |
| --- | --- |
| *Dependent variable* |  |
| BMI | Body mass index in kg/m<sup>2</sup> |
| *Independent variables* |  |
| Business closure | 1=business closed between waves; 0=still working for previous wave employer |
| BMI mPGS | Body mass index mean polygenic risk score, standardized |
| BMI vPGS | Body mass index variance polygenic risk score, standardized |
| Female gender | 1=female; 0=male |
| Age | Age in years |
| Married | 1=Married/partnered; 0=Divorced/separated/widowed/single |
| Highest degree obtained | Binary (0/1) variables for no degree or high school degree. The omitted category is associate's/bachelor's/professional degree. |
| Region dummies | Binary (0/1) variables for Census region of residence: Northeast (New England and Mid Atlantic); Midwest (EN Central and WN Central); South (S Atlantic, ES Central, and WS Central). The omitted category is West (Mountain and Pacific). |
| Household income (log) | Log of total (respondent + spouse) household income in 2010 dollars. Includes earnings, household capital income, income from all pensions and annuities, income from social security disability and supplemental social security income; income from social security retirement, spouse or widow benefits, income from unemployment or workers compensation, income from veteran's benefits, welfare and food stamps, alimony, other income, and lump sums from insurance, pension and inheritance. |
| Household wealth (\$100k) | Total household income in 2010 dollars divided by 100,000 for scalar consistency. It is the sum of the value of primary residence, net value of real estate (not including primary residence), net value of vehicles, net value of businesses, net value of stocks, mutual funds, and investment trusts, value of checking, savings or money market accounts, value of CD, government savings bonds, and T-bills, net value of bonds and bond funds, and the net value of all other savings, less the value of all mortgages/land contracts (primary residence), value of other home loans (primary residence), and the value of any other debt. |
| Firm size<sup>a</sup> | Binary (0/1) variables for firm size categories: Less than or equal to 4 employees; 5-14 employees; 15-24 employees; 25-99 employees; 100-499 employees. The omitted category is firm size greater than or equal to 500 employees. |
| Part time | 1=works part time; 0=does not work part time. |
| Industry<sup>a</sup> | Binary (0/1) variables for industry categories: agriculture, fishing, or farming; construction or mining; manufacturing; trade; public services; finance, insurance, or real estate; public administration. The omitted category is miscellaneous services. |
| Occupational status<sup>a</sup> | Binary (0/1) variables for blue collar and service workers. The omitted category is white collar workers. |
| Job tenure<sup>a</sup> | Current job tenure in years. |
| Health status | 1=excellent or very good self-reported health; 0=good, fair, or poor self-reported health. |
| Health insurance | 1=covered by a federal or employer-sponsored health insurance program; 0=otherwise. |
| Exercise | 1=exercises vigorously three or more times per week; 0=otherwise. |
| Ever smoke cigarettes | 1=smoked 100 or more cigarettes in their lifetime; 0=otherwise. |
| Cigarettes per day<sup>a</sup> | Total number of cigarettes smoked per day, excluding pipes or cigars. Variable is set equal to zero if respondent does not smoke. |
| Drinks per week | Total number of alcoholic drinks per week |
| Doctor diagnosed psychiatric issue<sup>a</sup> | 1=reports doctor diagnosed emotional or psychiatric problems; 0=otherwise. |
| Survey year | Binary (0/1) variables for 1994-2012. The omitted year is 1992. |

<sup>a</sup>Variables with additional category for missing values. Analyses also controls for the first 10 principal components of the European ancestry genetic data.

**Supplementary Table 2.** Descriptive statistics for the analytic sample

| Variable | Mean (SD) or<br>n (%) | Min | Max |
| --- | --- | --- | --- |
| BMI | 27.747 (5.251) | 15.3 | 63.2 |
| BMI mPGS | 0 (1) | -3.06 | 3.228 |
| BMI vPGS | 0 (1) | -2.789 | 5.758 |
| Female | 6,620 (0.555) | 0 | 1 |
| Age | 57.158 (4.252) | 50 | 69 |
| Married or partnered | 9,677 (0.811) | 0 | 1 |
| *Education* |  |  |  |
| No degree | 721 (0.06) | 0 | 1 |
| High school degree | 6,229 (0.522) | 0 | 1 |
| College degree | 4,982 (0.418) | 0 | 1 |
| Household income (log) | 11.33 (0.852) | 0 | 15.763 |
| Household wealth (\$100k) | 3.627 (13.728) | -10.079 | 1098.5 |
| Works part time | 1,369 (0.115) | 0 | 1 |
| *Firm size* |  |  |  |
| Firm size<=4 | 323 (0.027) | 0 | 1 |
| Firm size 5-14 | 688 (0.058) | 0 | 1 |
| Firm size 15-24 | 327 (0.027) | 0 | 1 |
| Firm size 25-99 | 1,253 (0.105) | 0 | 1 |
| Firm size 100-499 | 2,233 (0.187) | 0 | 1 |
| Firm size>=500 | 6,888 (0.577) | 0 | 1 |
| Missing firm size info | 220 (0.018) | 0 | 1 |
| *Industry* |  |  |  |
| Agriculture/Fishing/Farming | 81 (0.007) | 0 | 1 |
| Construction/Mining | 394 (0.033) | 0 | 1 |
| Manufacturing | 1,966 (0.165) | 0 | 1 |
| Trade | 1,496 (0.125) | 0 | 1 |
| Public Services | 881 (0.074) | 0 | 1 |
| Finance/Insurance/Real Estate | 761 (0.064) | 0 | 1 |
| Public Administration | 776 (0.065) | 0 | 1 |
| Misc. Services | 5,521 (0.463) | 0 | 1 |
| Missing industry info | 56 (0.005) | 0 | 1 |
| *Occupational status* |  |  |  |
| White Collar | 8,483 (0.711) | 0 | 1 |
| Blue Collar | 2,154 (0.181) | 0 | 1 |
| Service | 1,048 (0.088) | 0 | 1 |
| Missing occupation info | 247 (0.021) | 0 | 1 |
| Job tenure | 15.135 (10.859) | 0 | 48.5 |
| Missing tenure info | 23 (0.002) | 0 | 1 |
| *Health status* |  |  |  |
| Health excellent/very good | 7,605 (0.637) | 0 | 1 |
| Health insurance | 9,685 (0.812) | 0 | 1 |
| Exercise 3+ times/week | 4,320 (0.362) | 0 | 1 |
| Missing exercise info | 41 (0.003) | 0 | 1 |
| Ever smoke cigarettes | 6,420 (0.54) | 0 | 1 |
| Cigarettes per day | 2.852 (7.804) | 0 | 80 |
| Missing cigarettes per day info | 10,024 (0.84) | 0 | 1 |
| Drinks per week | 2.351 (5.607) | 0 | 84 |
| Missing drinking | 2,500 (0.21) | 0 | 1 |
| CES-D | 0.761 (1.423) | 0 | 8 |
| Missing CES-D | 1,585 (0.133) | 0 | 1 |

*Note:* Analytic sample consists of white, non-Hispanic workers aged 50-70 who were not self-employed.

**Supplementary Table 3.** Event time study analysis of differences in BMI between treatment and control groups pre-and post-job loss

|  | **1** | **2** | **3** |
| --- | --- | --- | --- |
|  | Analytic sample | High vPGS | Low vPGS |
|  | Beta (SE)<br>*p*-value | Beta (SE)<br>*p*-value | Beta (SE)<br>*p*-value |
| t-6 | -0.116 (0.142)<br>0.412 | -0.246 (0.226)<br>0.275 | 0.0794 (0.183)<br>0.664 |
| t-4 | -0.063 (0.123)<br>0.611 | -0.226 (0.179)<br>0.207 | 0.117 (0.173)<br>0.497 |
| t-1 (reference category) | - | - | - |
| t | -0.122 (0.107)<br>0.254 | 0.0426 (0.144)<br>0.767 | -0.318 (0.150)<br>0.035 |
| t+2 | 0.075 (0.154)<br>0.626 | 0.037 (0.207)<br>0.856 | 0.044 (0.236)<br>0.853 |
| Observations | 12,784 | 6,388 | 6,396 |
| Treated observations | 919 | 448 | 468 |
| Control observations | 11,865 | 5,940 | 5,928 |
| R-squared | 0.884 | 0.886 | 0.89 |

Abbreviations: SE, standard error. Robust standard errors in parentheses. Each regression is run separately for the full analytic sample (Column 1), and then stratified by high versus low vPGS groups (Columns 2 and 3). vPGS groups were stratified at the median. Additional covariates are listed in Table 1 and defined in detail in Supplementary Table 1. Individuals in the treated and control groups can have multiple observations. Unique N(treated)=374; unique N(control)=3,564.

**Supplementary Table 4.**
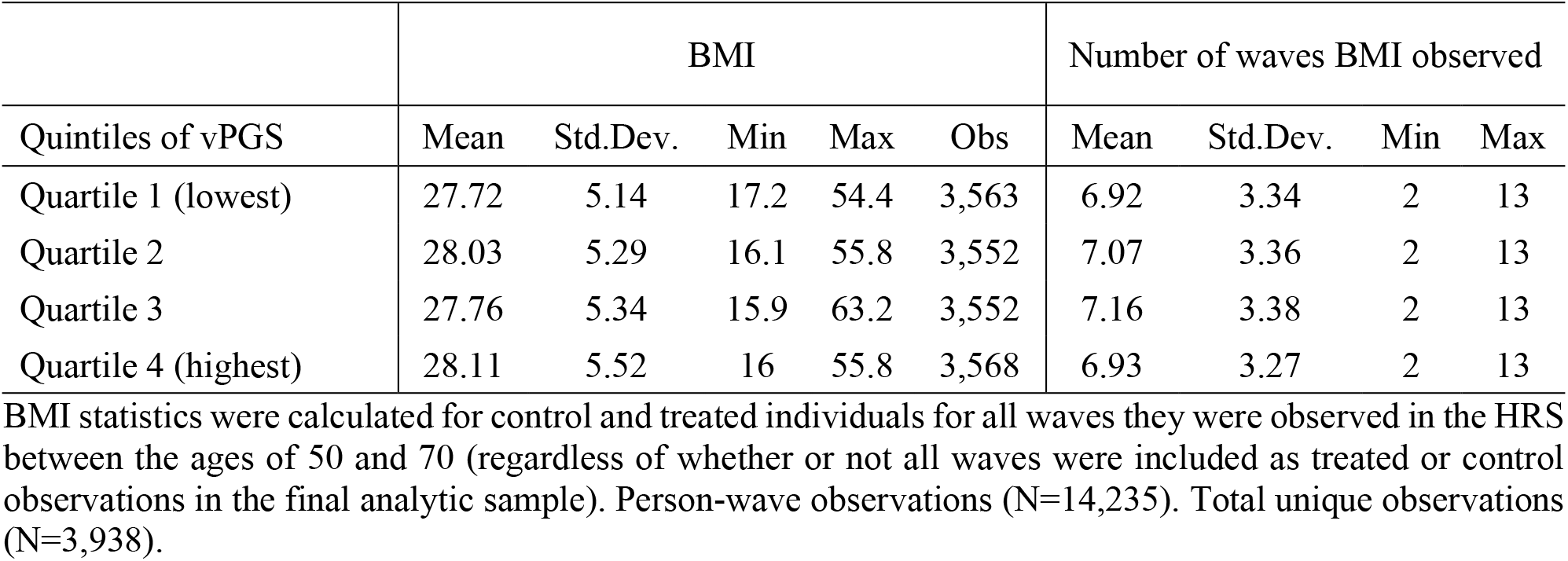
Mean within-person BMI by vPGS quartile

| Quintiles of vPGS | BMI |  |  |  |  | Number of waves BMI observed |  |  |  |
| --- | --- | --- | --- | --- | --- | --- | --- | --- | --- |
|  | Mean | Std.Dev. | Min | Max | Obs | Mean | Std.Dev. | Min | Max |
| Quartile 1 (lowest) | 27.72 | 5.14 | 17.2 | 54.4 | 3,563 | 6.92 | 3.34 | 2 | 13 |
| Quartile 2 | 28.03 | 5.29 | 16.1 | 55.8 | 3,552 | 7.07 | 3.36 | 2 | 13 |
| Quartile 3 | 27.76 | 5.34 | 15.9 | 63.2 | 3,552 | 7.16 | 3.38 | 2 | 13 |
| Quartile 4 (highest) | 28.11 | 5.52 | 16 | 55.8 | 3,568 | 6.93 | 3.27 | 2 | 13 |

BMI statistics were calculated for control and treated individuals for all waves they were observed in the HRS between the ages of 50 and 70 (regardless of whether or not all waves were included as treated or control observations in the final analytic sample). Person-wave observations (N=14,235). Total unique observations (N=3,938).

